# A High-resolution Haplotype-resolved Reference Panel Constructed from the China Kadoorie Biobank Study

**DOI:** 10.1101/2022.12.14.22283491

**Authors:** Canqing Yu, Xianmei Lan, Ye Tao, Yu Guo, Dianjianyi Sun, Puyi Qian, Yuwen Zhou, Robin Walters, Linxuan Li, Iona Millwood, Jingyu Zeng, Pei Pei, Ruidong Guo, Huaidong Du, Tao Yang, Ling Yang, Fan Yang, Yiping Chen, Fengzhen Chen, Xiaosen Jiang, Zhiqiang Ye, Fangyi Ren, Lanlan Dai, Xiaofeng Wei, Xun Xu, Huanming Yang, Jian Wang, Zhengming Chen, Huanhuan Zhu, Jun Lv, Xin Jin, Liming Li

## Abstract

Precision medicine relies on high-accuracy individual-level genotype data. However, the whole-genome sequencing (WGS) is currently not suitable for studies with very large sample sizes due to budget constraints. It is particularly important to construct highly accurate haplotype reference panel for genotype imputation. In this study, we selected 9,950 individuals from the China Kadoorie Biobank (CKB) cohort and 50 Chinese samples from the 1000 Genome Project (1KGP) for medium-depth WGS to construct a CKB reference panel. The results of imputing microarray datasets showed that the CKB panel outperformed the extended high coverage 1KGP, TOPMed, ChinaMAP, and NuyWa panels in terms of both the number of well-imputed variants and imputation accuracy. In addition, we have completed the imputation of over 100,000 CKB microarray data with the CKB panel, and the after-imputed genotype data is the hitherto largest whole genome data of the Chinese population. Finally, we developed an online server for offering free genotype imputation service based on the CKB reference panel (https://db.cngb.org/imputation/). We believe that the constructed CKB reference panel is of great value for imputing microarray or low-depth genotype data of Chinese population. The imputation-completed 100,000 microarray data are fundamental resources of population genetic studies for complex traits and diseases in the Chinese population.

## Introduction

In recent years, precision medicine has made remarkable achievements in complex diseases retreatment and development of target drugs by using molecular biological information (e.g., individual genome) and clinical symptoms [1, 2]. Precision medicine relies on high-throughput whole genome data to implement individual-based clinical diagnosis and treatment for patients. However, although the cost of whole genome sequencing (WGS) technology has been greatly reduced, there is still a budget problem for large-scale population research. Most researchers still prefer the low-cost microarray-based genotyping technology, which sequences known loci to obtain genotype data for follow-up analysis. But microarray cannot mine novel mutation sites related to the disease, so there are limitations in the interpretation of the genetic mechanism of the disease. At present, the common method is to impute the microarray data at the whole genome level based on the appropriate reference panel thus to obtain the whole genome data for a population. The selection of reference genome plays an important role in the imputation accuracy of genome data and subsequent analysis results.

Internationally, the haplotype map (HapMap) [3, 4], 1000 genome project (1KGP) [5, 6] the Haplotype reference consortium (HRC) [7], and the trans-omics for precision medicine (TOPMed) [8] have been launched. The HapMap project is the next major human genomic program after the International Human Genome Project. In 2007, the HapMap (phase 3) sequenced 1,184 individuals from 11 populations. In 2015, Chinese, British and American scientists jointly announced the completion of the Thousand Human Genome Project (phase 3), which sequenced the whole genomes of 2,504 individuals from 26 global populations and created the most comprehensive genetic polymorphism map of the human genome. The 1KGP panel is the most-commonly used genome data to date. Recently, the expanded 1KGP cohort including 602 trios were published, in which all 3,202 samples were sequenced to a high depth of 30X [6]. In 2016, the HRC project integrated 20 studies, such as UK10K and 1KGP, and created a reference panel with 32,470 individuals mostly with low-coverage WGS data [7]. The latest TOPMed reference panel collected 97,256 individuals, including 47,159 Europeans, 24,267 Africans, 17,085 admixed Americans, 1,184 East Asians, 644 South Asians, and other populations [8].

In recent years, in addition to the international haplotype reference projects, national haploid genome sequence consortiums have also been initiated in various countries, including Netherlands, Denmark, Iceland, and Singapore. The Dutch Human Genome Project sequenced 250 pedigrees at moderate depth (12X) to construct haploid reference sequences, substantially improving the accuracy of genotype inference for low-frequency variants [9]. The Danish Genome Project sequenced 50 Danish families at high-depth (80X) WGS to construct the first Danish genome-wide high-precision haplotype reference panel [10]. The Icelandic Genome performed high-depth (20X) WGS on approximately 2,000 individuals to create haplotype reference sequences, significantly improving the efficacy of association analysis and complex disease studies [11]. The SG10K reference panel sequenced 4,810 individuals, including 2,780 Chinese, 903 Malays, and 1,127 Indians, with an average sequencing depth of 13.7X [12]. This database is a valuable resource to advance the genetic study of complex traits and diseases in Asians.

China has the largest population in the world, producing a huge amount of genetic resources, and should make a greater contribution to human genetics and complex disease research. However, the lack of high-quality haplotype reference sequences has become a bottleneck in the fields of population genetics and molecular biology. Fortunately, in the past two years, researchers have constructed reference panels based on Chinese population: the ChinaMAP (China Metabolic Analytics Project) and the Nyuwa reference panels. The ChinaMAP consortium performed 40X deep WGS on 10,588 individuals collected from different regions and ethnicities in China [13, 14]. The library construction and WGS were performed on the BGISEQ-500 platform at BGI-Genomics. The ChinaMAP reference panel is a high-quality genetic variation database of Chinese population and plays an essential role in the analysis of Chinese population structure, genetic variation spectrum, and pathogenic variants. The NyuWa reference panel includes 2,999 high-depth (26.2X) WGS samples collected from 23 administrative regions of China [15]. It is important to expand the diversity of genetic resources and improve the accuracy of medical research in Chinese population.

The China Kadoorie Biobank (CKB), previously known as the Kadoorie Study of Chronic Disease in China (KSCDC), is an international collaborative research project on chronic diseases jointly conducted by Peking University, Chinese Academy of Medical Sciences and University of Oxford, UK [16]. It is a gargantuan prospective study and the largest Chinese population cohort to date. During 2004-2008, over 510,000 adults were recruited from ten geographically defined regions in China. The study aims to establish a database of blood samples and clinical information and to investigate the main genetic and environmental causes of common chronic diseases. To date, the CKB cohort has achieved numerous influential findings in clinical studies, such as the relationship between smoking, physical activity, fresh fruit intake, egg consumption and the risk of cardiovascular disease [17-20], the association between diabetes and the risk of death [21], and the relationship between smoking, alcohol and tea consumption and esophageal cancer [22]. However, unfortunately, there are no large-scale population genetics and genetic background studies of complex traits and diseases based on the CKB cohort [23]. A major reason for this is the lack of high-density genetic data. Although microarray testing (Affymetrix Axiom myDesign) of over 100,000 samples has been completed, the data are still not comparable to WGS data in terms of the number of genetic variants and the detection of novel loci.

In this work, we constructed a high-resolution haplotype-resolved reference panel based on 9,950 individuals from the CKB cohort and 50 Chinese samples from the 1KGP study, with an average sequencing depth of 15X. We evaluated the imputation performance of the CKB reference panel from the perspective of number of imputed variants and imputation accuracy. The compared reference panels include the extended high coverage 1KGP, the newly developed TOPMed, the ChinaMAP and the NyuWa panels built from the Chinese population. In addition, based on the constructed CKB panel, we completed the genotype imputation for over 100,000 microarray samples and obtained the largest whole genome data in the Chinese population. We also created an online imputation server to offer free genotype imputation service.

## Results

### Data quality

Following the standard genotype analysis procedures, we obtained that the mean sequencing depth of the 10,000 samples in the CKB reference panel was 15.55X (Figure S1a). The inferred sample sex suggested that 5,572 (56%) individuals were female and 4,423 (44%) individuals were male (Figure S1b), which was highly consistent with the sex distribution of entire CKB cohort (i.e., female: 59.0%, male: 41.0%). We then removed 27 samples with abnormal GC content (GC<40% or GC>44%) (Figure S1c). Note that, during the construction of the CKB reference panel, we did not remove related samples since the genetic relatedness can be modeled and improve haplotype phase accuracy. In the end, we removed 9 genetically related samples from our panel as the related samples can distort the population allele frequency estimation in the subsequent analysis. Thus, the final reference panel comprised of 9,964 samples and 129.74M variants, including 113.73M SNPs and 16.01M INDELs for 22 autosomes.

A comprehensive comparison of the CKB reference panel and other panels in terms of sample size, sequencing depth, and imputed variants was provided in Table 1. Specifically, the ChinaMAP reference panel, which contained the largest sample of 10,588 with the deepest WGS sequencing (40.83X), included 136.75M SNPs and 10.70M INDELs. The NyuWa reference panel contained 71.06M SNPs and 8.19M INDELs. The extended 1KGP panel consisted of 111.05M SNPs and 14.44M INDELs. The TOPMed panel consisted of 286.07M SNPs and 22.04M INDELs. By contrast, the CKB and ChinaMAP reference panels were the only two panels composing of over a 100 million variants in Chinese population.

**Table 1.** The information of CKB and other reference panels.

| Reference panel | Sample size | Sequencing depth | Variants | SNP | INDEL | Ancestries |
| --- | --- | --- | --- | --- | --- | --- |
| **CKB** | 10,000 | 15.55X | 129,743,542 | 113,731,044 | 16,012,498 | Chinese |
| **ChinaMAP** | 10,588 | 40.8X | 147,448,941 | 136,745,826 | 10,703,115 | Chinese |
| **Extended 1KGP** | 3,202 | 34X | 125,484,020 | 111,048,944 | 14,435,076 | Multiple ancestries |
| **TOPMed** | 97,256 | >30X | 308,107,085 | 286,068,980 | 22,038,105 | Multiple ancestries |
| **NyuWa** | 2,999 | 26.2X | 79,226,351 | 71,057,961 | 8,168,390 | Chinese |

In addition, we also calculated three quality indicators for SNPs: the heterozygous/homozygous (het/hom) ratio, the transition/transversion (Ti/Tv) ratio, and the non-reference genotype concordance rate (NRC). The het/hom ratio is highly dependent on ancestry and the median value for Asians is around 1.4 [24]. The Ti/Tv ratio reflected the quality of SNP calling and the expected ratio would be close to 2.0 for human WGS data [25]. For the CKB reference panel, we obtained a het/hom ratio of 1.31 and a Ti/Tv ratio of 1.97, indicating high-quality of genotypic data in the constructed panel. The NRC is genotype-aware recall (a.k.a. sensitivity = TP/(TP+FN)). We used the genotype data of 50 1KGP samples with high-depth sequencing as actual status and their SNP calls in the CKB panel as the predicted data. The NRC for these 50 samples were calculated before and after genotype phasing implemented by Beagle 5.2 [26]. The average NRC increased from 0.9811 to 0.9927 and the improvement is more significant for samples with lower sequencing depth (Figure S1d).

### Novel variants and functional annotation

We defined novel variants that were not assigned a unique variant accession identifier (RS number) in dbSNP (Single Nucleotide Polymorphism Database, build 154) [27]. Thereby, the number of novel SNPs and INDELs are 50.16 million (44.1%) and 5.42 million (33.8%), respectively (Figure S1e, S1f). Note that, a site with different mutation variety compared to that in dbSNP (e.g., in panel: REF:ALT is A:-, while in dbSNP REF:ALT is CA:C) was also considered as a novel variant, which partially explained the relatively high proportion of novel sites [28, 29]. As expected, the vast majority of novel SNPs (99.99%) and INDELs (99.15%) were rare variants (MAF < 0.5%) (Figure S1e, S1f).

Based on the results of annotation analysis, 55% were intronic variants and 26% variants located in the intergenic region. The subsequent categories were non-coding variants (15%), upstream/downstream transcript variants (12%), regulatory variants (4%), variants in mRNA untranslated regions (1%), functional variants (0.8%), transcription factor binding sites (0.3%), and splice-site variants (0.1%) (Figure 2a). Among the functional variants, the most numerous class is missense mutations (Figure 2a). The SIFT and PolyPhen algorithms provided consistent prediction of deleterious variants, that was a large fraction (96%) were very rare variants (AC<=2) (Figure 2b). Over 72% variants can be predicted as deleterious by both algorithms (Figure 2c). For the low-frequency and common variants (MAF>0.005), 23 (0.3%) of them were annotated as deleterious. In particular, 7 variants were predicted as deleterious mutations by both SIFT and PolyPhen algorithms, 7 variants were uniquely annotated by SIFT, and 9 were uniquely annotated by PolyPhen (Table S2).

**Figure 1:**
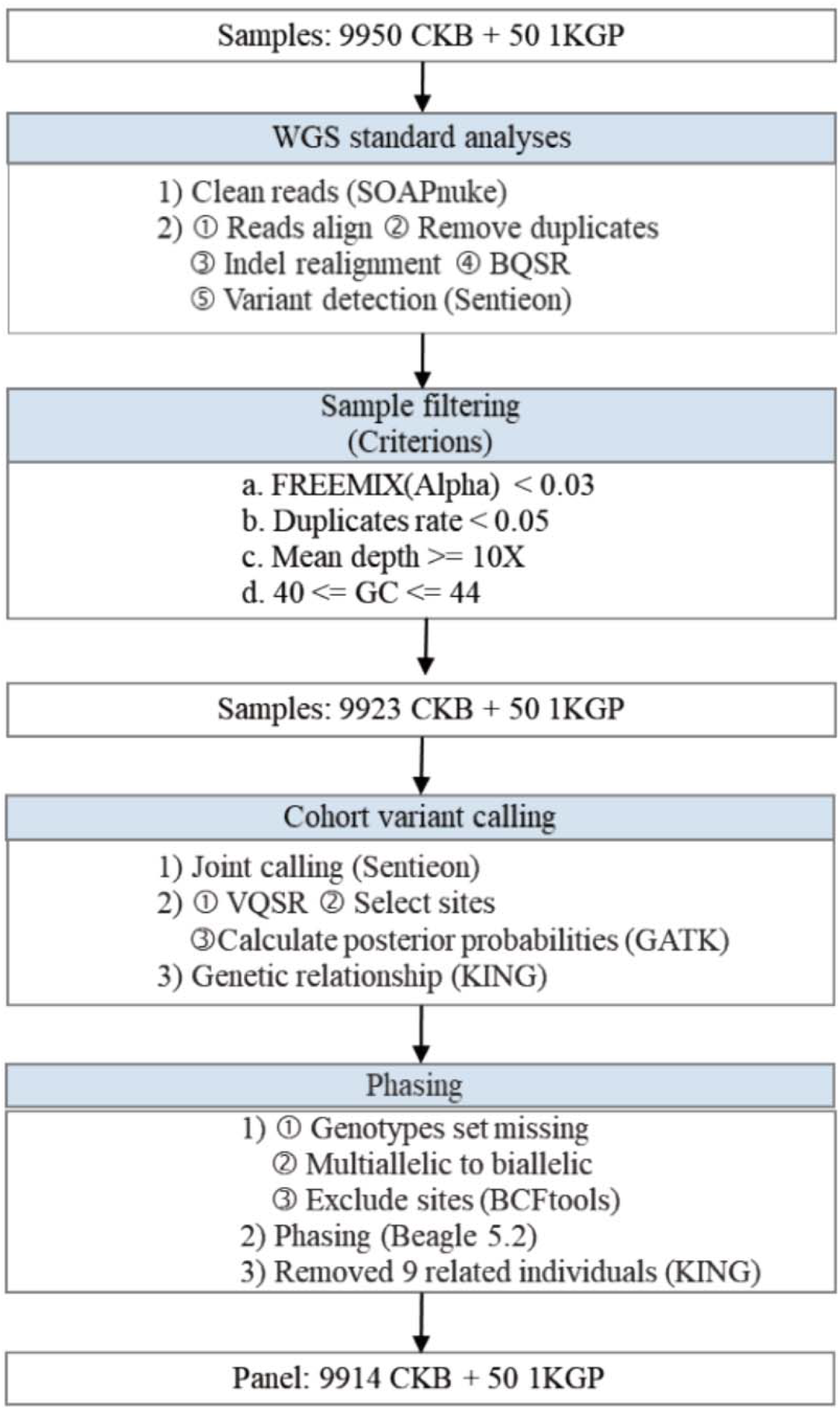
The workflow of panel construction.

**Figure 2:**
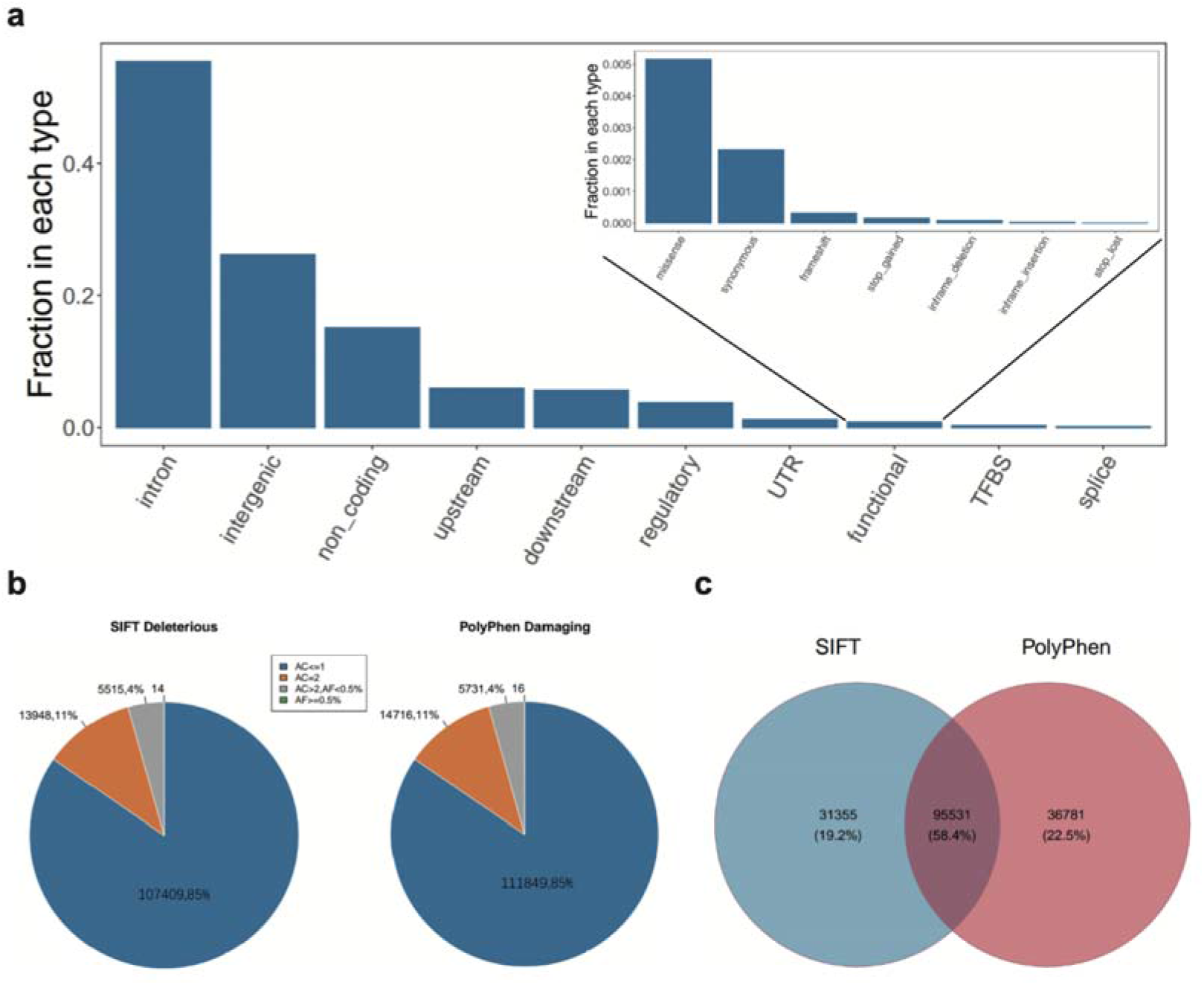
The annotations of the CKB reference panel. ***Notes*: a**. Annotations of all novel variants. The major bar chart showed all the 10 categories. The minor bar chart displayed the functional variants with more detailed roles. **b**. Distribution of novel deleterious variants annotated by SIFT (left) and damaging variants annotated by PolyPhen (right) for different allele counts or frequencies. **c**. The Venn diagram of numbers of novel deleterious variants annotated by SIFT and PolyPhen.

### Imputation performance evaluation

We compared the imputation performance of the CKB reference panel with that of the extended 1KGP [6], TOPMed [30], ChinaMAP [14], and NyuWa [31] from the perspective of number of imputed variants and imputation accuracy. We used 50 CKB and 50 1KGP microarray datasets as input samples to be imputed. The corresponding high-depth WGS data were used as ground truth datasets. In imputation of the CKB array data, the CKB reference panel provided the highest number of medium-quality imputed variants (10.86M), followed by the extended 1KGP (10.01M), NyuWa (9.23M), TOPMed (8.80M), and ChinaMAP (7.98M) reference panels. When focusing on only high-quality imputed variants, we observed that the ChinaMAP reference panel had the greatest percentage of high-quality variants (86.23%), followed by CKB (84.63%), TOPMed (80.22%), extended 1KGP (78.50%) and NyuWa (77.32%) (Figure 3a; Table 2). We note that the reason why the ChinaMAP provides the smallest number of medium-quality variants is that it automatically filters out almost half of low-quality variants in the actually used panel.

**Table 2.**
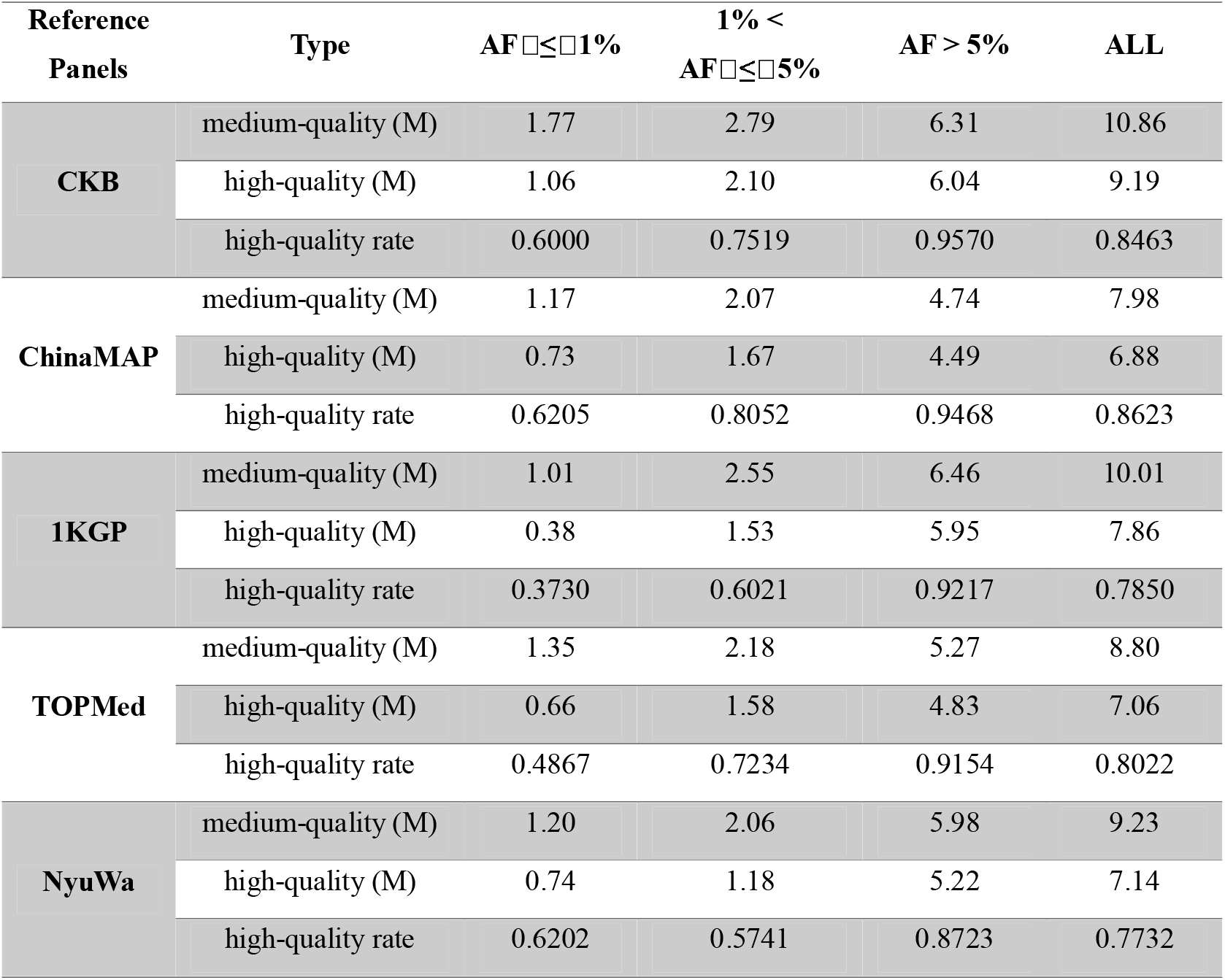
The high-quality and medium-quality imputed variants for imputing 50 microarray samples.

| Reference<br>Panels | Type | AF $\leq 1\%$ | 1% <<br>AF $\leq 5\%$ | AF > 5% | ALL |
| --- | --- | --- | --- | --- | --- |
| **CKB** | medium-quality (M) | 1.77 | 2.79 | 6.31 | 10.86 |
|  | high-quality (M) | 1.06 | 2.10 | 6.04 | 9.19 |
|  | high-quality rate | 0.6000 | 0.7519 | 0.9570 | 0.8463 |
| **ChinaMAP** | medium-quality (M) | 1.17 | 2.07 | 4.74 | 7.98 |
|  | high-quality (M) | 0.73 | 1.67 | 4.49 | 6.88 |
|  | high-quality rate | 0.6205 | 0.8052 | 0.9468 | 0.8623 |
| **1KGP** | medium-quality (M) | 1.01 | 2.55 | 6.46 | 10.01 |
|  | high-quality (M) | 0.38 | 1.53 | 5.95 | 7.86 |
|  | high-quality rate | 0.3730 | 0.6021 | 0.9217 | 0.7850 |
| **TOPMed** | medium-quality (M) | 1.35 | 2.18 | 5.27 | 8.80 |
|  | high-quality (M) | 0.66 | 1.58 | 4.83 | 7.06 |
|  | high-quality rate | 0.4867 | 0.7234 | 0.9154 | 0.8022 |
| **NyuWa** | medium-quality (M) | 1.20 | 2.06 | 5.98 | 9.23 |
|  | high-quality (M) | 0.74 | 1.18 | 5.22 | 7.14 |
|  | high-quality rate | 0.6202 | 0.5741 | 0.8723 | 0.7732 |

**Figure 3:**
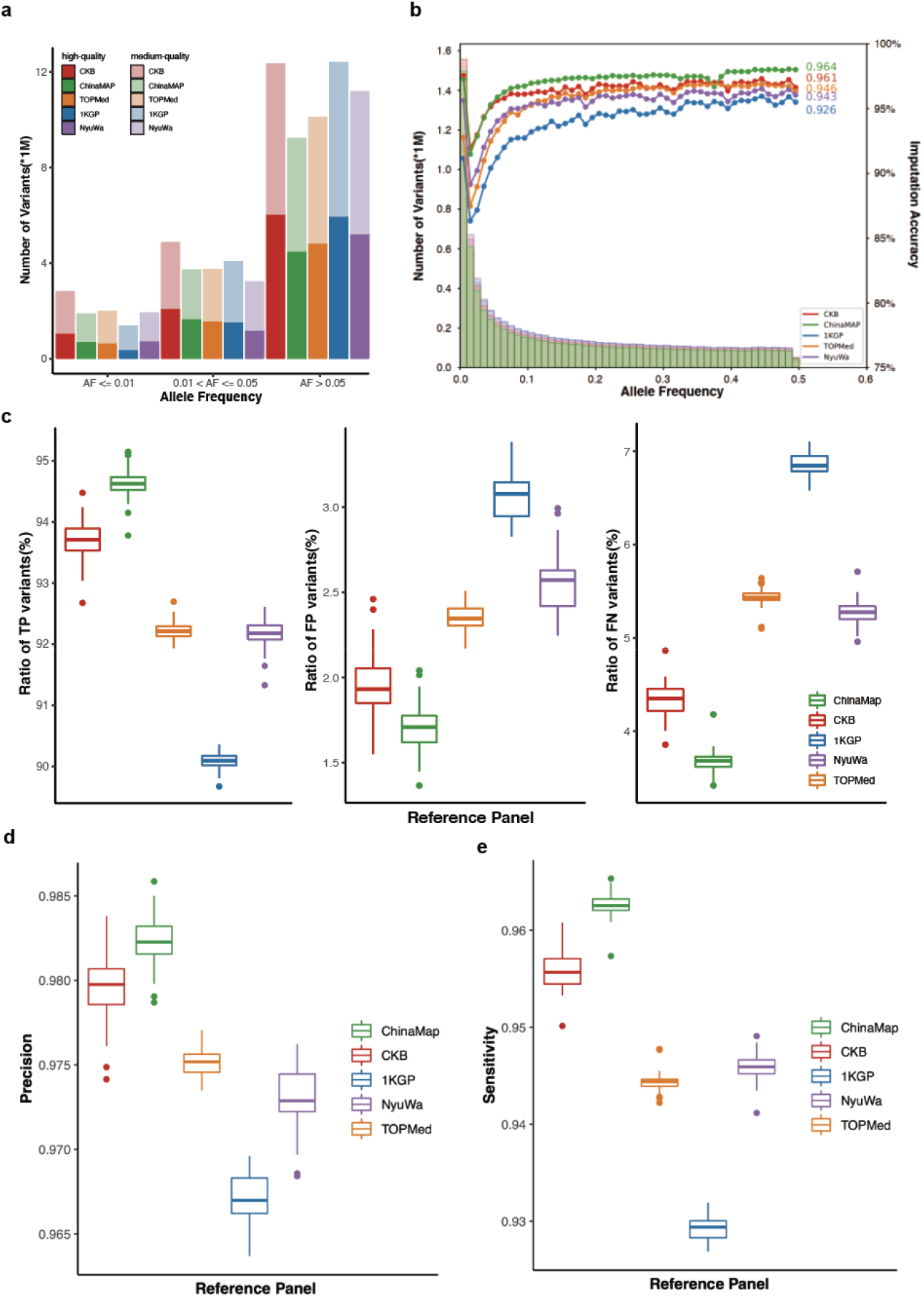
The performance for imputing 50 CKB microarray data. ***Notes*: a**. The numbers of high- and medium-quality imputed variants under different AF (allele frequency) by using different reference panels. **b**. The histogram of imputed variants and Pearson correlation coefficients for different panels. **c**. The boxplots of the ratios of true positive (TP), false negative (FN), and false positive (FP) variants. **d, e**. The imputation precision **(d)** and sensitivity **(e)** of the reference panels.

We evaluated the imputation accuracy by using three measurements: Pearson correlation coefficient (R^2^), precision, and sensitivity. The mean R^2^ of the compared reference panels were 0.964 (ChinaMAP), 0.961 (CKB), 0.946 (TOPMed), 0.943 (NyuWa), and 0.926 (extended 1KGP) (Figure 3b). For the ratios of TP, FP, and FN variants, the ChinaMAP reached the highest ratio of TP variants (94.62%), subsequently followed by CKB (93.71%), then followed by TOPMed (92.21%), NyuWa (92.18%), and extended 1KGP (90.09%). Meanwhile, the ChinaMAP obtained the lowest ratios of FP (1.71%) and FN (3.68%) variants, and for the CKB panel, the two ratios were 1.93% and 4.35%, respectively. These ratios in TOPMed (FP: 2.35% and FN: 5.43%) and NyuWa (FP: 2.57% and FN: 5.28%) were slightly higher than those in the CKB panel. The extended 1KGP reference panel had the highest ratio of FP (3.08%) and FN (6.85%) variants (Figure 3c). Consequently, the ChinaMAP attained the highest precision of 98.23%, followed by CKB (97.98%), TOPMed (97.52%), NyuWa (97.29%), and extended 1KGP (96.70%). For sensitivity, the ChinaMAP and CKB panels reached 96.25% and 95.57%, respectively. Following that, the NyuWa, TOPMed, and extended 1KGP obtained the sensitivity of 94.59%, 94.44%, and 92.94%, respectively. The CKB reference panel achieved very similar R^2^, precision, and sensitivity compared to the ChinaMAP, displaying an outstanding imputation performance (Figure 3d, 3e).

In the imputation of the 1KGP array data, we compared the performance of the CKB panel with that of ChinaMAP and TOPMed. We excluded the extended 1KGP panel as it had overlap samples with the array data, and also excluded NyuWa panel as the web server is unstable and not accessible for submitting jobs currently. The CKB reference panel provided the highest number of medium-quality imputed variants (9.75M), followed by TOPMed (7.83M) and ChinaMAP (6.98M) reference panels. When focusing on only high-quality imputed variants, we observed that the ChinaMAP reference panel had the greatest percentage of high-quality imputed variants (87.92%), followed by TOPMed (84.46%) and CKB (84.11%) (Figure S2a). For the Pearson correlation coefficient R^2^, both the CKB and ChinaMAP panels achieved 0.979, while the TOPMed had a lower R^2^ of 0.965 (Figure S2b).

### Imputation of over 100,000 microarray data

The imputation-completed whole genome data contained 42.61M medium-quality variants and 17.45M high-quality variants. To assess the imputation performance on over 100,000 CKB microarray data, we calculated the Pearson correlation coefficients (R^2^) of 50 CKB samples with imputed genotype and high-depth WGS data. Note that, we did not have WGS data for the 100,706 microarray data, thus could not use all of the samples as the true set. As an alternative, we used a subset of 50 individuals with WGS data as samples being evaluated. Consequently, the averaged R^2^ was 0.972. Remember that when we simulated only these 50 microarray samples, the averaged R^2^ was 0.961 (Figure 4). This phenomenon verified that the size of to-be-imputed samples also partly contributed to the imputation results. The high imputation accuracy of 0.972 demonstrated that the proposed CKB reference panel is quite capable of imputing extensive data.

**Figure 4:**
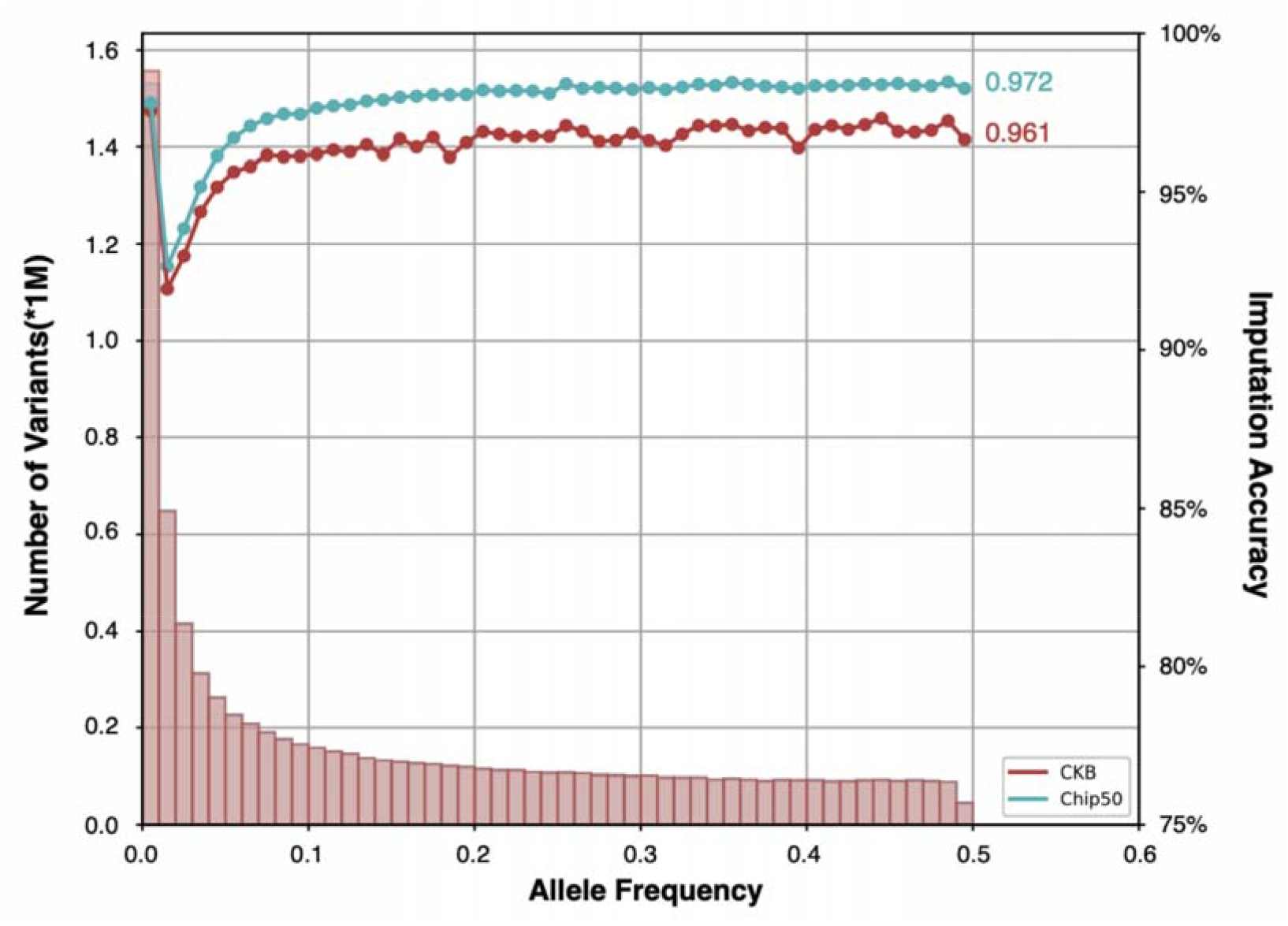
The Pearson correlation coefficient for imputing over 100,000 samples. ***Notes***: The Pearson correlation coefficient was calculated between the true high coverage WGS data and 50 microarray data either extracted from the entire 100,706 after-imputed dataset (green) or imputed alone (red).

### GWAS analysis of simulated data

For the PCA analysis of individuals from the CKB panel, a total of 1,340,477 biallelic SNPs were used. The first PC represents the south-north geographical information of China (Figure S3). With imputed phenotype data under the null hypothesis that there were no associated SNPs, the GWAS analysis did not identify any significant signals, and the p-values were uniformly distributed (Figure S4a, S4b) as expected. When the phenotype was generated by involving the effects of SNPs, the GWAS study successfully discovered causal SNPs and those in high linkage disequilibrium (LD) (Figure S5). Specifically, in addition to the five randomly selected causal SNPs (rs3003378, rs6764623, rs10905649, rs13254191, and rs10915307), high-LD SNPs (e.g., rs12564681, rs11923809, rs7092291, rs545854, rs12123277) were also identified. The results of GWAS analysis with simulated data under both null and alternative hypotheses demonstrated the high-quality of genotype data.

## Methods

### Subjects

In this project, we constructed a reference panel based on 9,950 samples from the CKB cohort and 50 samples from the 1KGP project. The CKB project recruited over 510,000 adults aged from 30 to 79 in ten (five urban, five rural) geographic regions of China. These 9,950 individuals were stroke cases from the cohort. The 50 1KGP samples included 20 Han Chinese and 30 Southern Han Chinese. Written informed consent was obtained from all participants from the CKB cohort.

### DNA samples and library construction

The whole genome sequencing (WGS) was performed for the 10,000 samples. Specifically, DNA concentration was measured by ExKubit dsDNA HS Assay Kits (Shanghai ExCell Biology, Inc) and Fluostar Omega Microplate Reader (BMG Labtech GmbH). The DNA quality was evaluated by agarose gel electrophoresis at a constant voltage (180V) for 35 minutes. The DNA shearing was done by the Covaris E220 ultrasonics DNA shearing instruments. The DNA purification and fragment size selection were applied by VAHTS DNA Clean Beads (Vazyme, #N411). The libraries were constructed on BGI’s DNBseq-T10×4RS platform and the loading DNA concentration was above 12 ng/μl. The paired-end 100bp (PE100) WGS with 350bp insert sizes was performed on the MGI DNBSEQ sequencing platform. The target sequencing depth is 15X per sample, about 45 GB in size.

### Variant calling and sample quality control

To perform variant calling on each sample (a.k.a. individual variant calling), we first applied SOAPnuke (version 2.1.1; -n 0.1 -l 12 -M 2) [32] to filter low quality reads and remove adapter sequences. Then, we obtained aligned Binbary Alignment/Map (BAM) files by aligning sequence reads to the GRCh38 human reference genome assembly with Sentieon (version 202010.04) bwa-mem algorithm [33]. On the sorted and aligned BAM files, we used Sentieon drivers LocusCollector to collect information on duplicates and Dedup to remove the duplicates. For regions that contain insertions or deletions (INDELs), we further performed local realignment around INDELs to correct for mapping errors and increase the quality of INDEL detection by using the Sentieon Realigner algorithm. To increase the accuracy of variant calling, we carried out base quality score recalibration (BQSR) to BAM files based on the Sentieon QualCal algorithm, which created a recalibration table. This table file was then applied as an input to Sentieon Haplotyper for single nucleotide polymorphisms (SNPs) and INDELs detection. After all these steps, we obtained the called variant sites for each sample in gVCF format.

Before performing joint variant calling, we first selected samples with (1) no evidence of contamination (VerifyBamID FREEMIX < 0.03) [34]; (2) high library quality measured by reads duplication rate < 0.05; (3) mean sequencing depth >= 10X; and (4) GC content between 40 and 44. We used SAMtools 1.3 [35] idxstats tool to infer sex by calculating coverage ratio of X-chromosome and autosomes. Samples with X-chromosome/autosomes ratio above 0.7 were inferred as female, otherwise male.

The joint variant calling was then performed by GVCFtyper algorithm implemented in Sentieon, followed by variant quality score recalibration (VQSR) for SNPs and INDels separately using GATK [25]. In this way, we first built the models with VariantRecalibrator and then applied it in ApplyVQSR. After that, ExcessHet > 54.69 and low-quality sites that did not pass VQSR were filtered out by SelectVariants. Finally, we calculated genotype posterior probabilities by CalculateGenotypePosteriors.

### Reference panel construction

After calculated genotype posterior probabilities, we further set low quality genotypes (GQ < 20) to missing and then removed sites with minimum count <1 or with missing alternate (ALT) allele. We also split a multiallelic row with more than one ALT allele to biallelic rows, with each ALT allele in a separate row. Next, we performed genotype phasing (a.k.a. phasing/haplotype estimation), which is the process of statistical estimation of haplotypes from genotype data. This step was done by Beagle 5.2 [36]. To build our reference panel, we made the final step of removing close relatives up to the second degree. The genetic relationship was generated by KING 2.2.7 [37]. After then, we obtained the final reference panel, which we named as CKB reference panel. The workflow of construction process was provided in Figure 1. We annotated all variants in the panel with the Ensembl Variant Effect Predictor (VEP) [38] by using plug-ins SIFT [39] and PolyPhen [40] algorithms.

### Evaluation of the imputation performance

We conducted extensive scenarios to evaluate the imputation performance of the CKB panel and others, including the extended 1KGP, TOPMed, ChinaMAP, and NyuWa reference panels. There were two datasets to be imputed: the CKB and 1KGP microarray data. From the CKB cohort, 50 randomly selected samples independent of that in the CKB reference panel were genotyped in both SNP array and high coverage WGS (30X). The 50 1KGP microarray samples were all Chinese and also independent of those in the CKB reference panel. To evaluate the imputation performance, we compared number of imputed variants and imputation accuracy. For the imputed variants, we defined high-quality variants with imputed info-score ≥ 0.8 and medium-quality variants with imputed info-score ≥ 0.4. For the imputation accuracy, we calculated Pearson correlation coefficient (R^2^), precision, and sensitivity. The 30X WGS data were treated as ground truth when computing imputation accuracy between the imputed and true genotypes. For the CKB and extended 1KGP panels, we performed imputation procedures locally; while for TOPMed, ChinaMAP, and NyuWa, we submitted jobs to their online imputation servers and downloaded the after-imputed files.

To assess the precision and sensitivity, we first calculated the true positive (TP), false positive (FP), false negative (FN), and true negative (TN). The TP indicates that the imputed genotype correctly predicts the true WGS genotype. The FP is an error classification where the imputed genotype incorrectly indicates the presence of a WGS variant. The FN is also an error classification where the imputed genotype incorrectly indicates the absence of a WGS variant. The TN is an outcome where the predicted genotype correctly predicts the case of homozygous reference calls. The details of 3×3 confusion matrix of defining TP, FP, FN, and TN were provided in Supplementary Table S1. To eliminate the bias caused by the number of imputed variants, we compared the ratios of TP, FP, and FN instead of their counts directly. The TN value for all panels was zero. The ratio of TP was calculated by TP/(TP+FP+FN+TN), same for FP and FN. The precision was computed by TP/(TP+FP) and the sensitivity was computed by TP/(TP+FN).

### Imputation for over 100,000 microarray data

We imputed 100,706 CKB microarray data based on the CKB reference panel in Beagle 5.2 [36]. Note that, the 50 samples with both microarray and high coverage WGS data were included in the 100,706 individuals. In order to do imputation efficiently, we randomly divided the 100,706 samples into 21 chunks, in which 20 chunks contained 4,800 samples and 1 chunk contained 4,706 samples. Then, we parallelly executed genotype imputation for these chunks. To assess the performance for imputing such a large volume of data using the developed CKB panel, we extracted the 50 after-imputed microarray samples and calculated the Pearson correlation coefficients with their high coverage WGS set. We also compared this imputation accuracy with that of imputing the 50 microarray samples alone.

### PCA analysis for the CKB reference panel

To detect population stratification, we carried out principal component analysis (PCA) [41, 42] of genotype data in the CKB reference panel. The PCA analysis was carried out in Plink 1.9 [43] with autosomal biallelic SNPs satisfying the following conditions (1) MAF ≥ 1%, (2) genotyping rate ≥ 90%, (3) Hardy-Weinberg-Equilibrium (HWE) P-value > 1E-06, and (4) low LD (r2 < 0.5) with other variants in windows of 50 SNPs with steps of 5 SNPs.

### GWAS analysis of simulated data

In this section, we aimed to perform genome-wide association study (GWAS) of simulated phenotypic values, while the genotype data was a combination of the CKB reference panel and after-imputed 100,706 microarray data. First, we performed PCA analysis of genotype data by using PCAone (https://github.com/Zilong-Li/PCAone), which was applicable for large samples. Then, we simulated phenotype data under null and alternative hypotheses, separately. Under the null hypothesis that none of the SNPs were associated with the phenotype, we generated a vector of phenotypic values from a standard normal distribution. Under the alternative hypothesis that the phenotype data was generated from a linear regression model by using five SNPs as independent variables with randomly assigned effects size β. The causal SNPs included rs3003378 (β=0.02), rs6764623 (β=0.01), rs10905649 (β=0.02), rs13254191 (β=0.03), and rs10915307 (β=0.01). We used PC1 to PC5 and sex of the participants as the covariates to do GWAS analysis in Plink 2.0 [44]. We reported Manhattan plots, QQ-plots, histograms, and regional plot for the GWAS results.

### Online imputation service

We developed an online imputation server to offer genotype imputation service, which allows users to run imputation tasks free and safely in an easy way. On the online server, we provided the CKB and 1KGP as available reference panels, GRch37 (hg19) and GRch38 (hg38) as human genome assembly, Minimac 4 [45, 46] and Beagle 5.2 [26] as imputation tools, and different population options (e.g., East Asian, European, African). Users can access the server via https://db.cngb.org/imputation/.

## Discussion

A population-specific haplotype reference panel is a collection of ancestral chromosome sequences that represents the genetic diversity of the population. High-precision reference panel is the basis for population genetic analysis and precision medicine. China has the largest population in the world and possesses vast amounts of genetic resources, but lacking high-quality reference panel has hindered the development of genetic studies and their application in human diseases based on the Chinese population. Fortunately, in the last two years, a few reference panels have been constructed for accurate genotype imputation in Chinese population, including the ChinaMAP and NyuWa.

In this work, we developed a high-resolution haplotype-resolved reference panel of 10,000 sequenced individuals from the CKB cohort and the 1KGP database. Even with medium sequencing depth (15.55X), the proposed CKB panel can compete with the ChinaMAP (40.80X) and outperform the extended 1KGP, TOPMed, and NyuWa in imputation accuracy measured by Pearson correlation coefficient, precision, and sensitivity. From the perspective of number of well-imputed variants, the CKB provided the largest number of medium-quality variants with info-score ≥ 0.4; for high-quality variants with info-score ≥ 0.8, the CKB panel obtained the second largest amount among all considered panels. What’s more valuable is that we completed the genotype imputation for over 100,000 CKB microarray data based on the constructed panel. The imputation accuracy reached as high as 0.972 and GWAS analysis based on the simulated data demonstrated the reliability of the extensive imputed data. This imputed dataset is the largest whole genome data for Chinese population to date and will certainly play a fundamental role in personalized medicine and drug development.

The ultimate goal of imputing genotype data is to increase statistical power of genetic association studies for identifying trait-associated SNPs and further to reveal the etiology of complex diseases. As the hitherto largest cohort of Chinese population, CKB collected abundant clinical data, including demographic, anthropometric, biochemical, radiographic traits, metabolomic tests, and diseases coded by ICD10 (international classification of diseases, version 10). There are over 1,500 diseases, mostly chronic diseases, such as heart attack, stroke, diabetes, cancers, and so on. As a significant future work, we will aim to perform GWAS analysis for the vast wealth of phenotypes and 100,000 imputed WGS genotype data. To the best of our knowledge, this should be the largest population genetic study in Chinese population and is also comparable to numerous international genomics research projects, for example, the UK Biobank study (https://www.ukbiobank.ac.uk/), the All of Us research program (https://allofus.nih.gov/), and the biobank Japan project (https://biobankjp.org/en/).

Most of the reference panels are now packaged into online imputation servers, such as the Michigan imputation server [46], TOPMed imputation server [46], ChinaMAP imputation server, NyuWa server, and our developed CKB imputation server. These imputation servers all provide free genotype imputation service by uploading to-be-imputed files and selecting reference panel, population, and imputation software. All the imputation results can be downloaded directly by clicking on filenames. Even though the online server provides a convenient way to impute genotype data, it typically cannot handle large-sized files, which causes difficulties in imputing large-sample data. When imputing large scale datasets, the individual-level reference panels are needed for offline imputation. Since the completion of the first human genome project in 2003 (https://www.genome.gov/human-genome-project), the only database that is fully publicly available is the 1000 Genome database. Sharing genomic data is critical for research efficiency, translating research results into products, and ultimately improving public health. Hence, we appeal the sharing of genomic and health related data with controlled management.

## Data Availability

The CKB reference panel and the after-imputed over 100,000 CKB microarray data have been deposited into CNGB Sequence Archive (CNSA) of China National GeneBank DataBase (CNGBdb) with accession number CNP0003405. All genotype data are shared with controlled management.All data produced are available online at

https://db.cngb.org/cnsa/

## Author contribution

Liming Li, Xin Jin, Jun Lv and Huanhuan Zhu conceived the study, designed the research program and managed the project.

Canqing Yu, Yu Guo, DianJianyi Sun, Zhiqiang Ye, Lanlan Dai, Fangyi Ren, and Puyi Qian finished the laboratory processing and data acquisition.

Canqing Yu, Xianmei Lan, Ye Tao, Yu Guo, Dianjianyi Sun, Puyi Qian, and Yuwen Zhou preprocessed the data, finished the quality control, and constructed the haplotype reference panel.

Ye Tao, Xianmei Lan, Linxuan Li, Pei Pei, Jingyu Zeng, Ruidong Guo, and Xiaosen Jiang finished the evaluation of the haplotype reference panel.

Robin Walters, Iona Millwood, Huaidong Du, Ling Yang, Yiping Chen, and Zhengming Chen provided useful advice on constructing and evaluating the reference panel.

Tao Yang, Fan Yang, Fengzhen Chen, Xiaofeng Wei built the online imputation server. Xianmei Lan, Ye Tao, and Huanhuan Zhu drafted the manuscript.

All authors participated in revising the manuscript.

## Acknowledgement

The most important acknowledgement is to the participants in the study and the members of the survey teams in each of the 10 regional centres, as well as to the project development and management teams based at Beijing, Oxford and the 10 regional centres.

This work was supported by grants (2016YFC0900500) from the National Key R&D Program of China, National Natural Science Foundation of China (32000398, 82192901, 82192904, 82192900), the China National GeneBank, Guangdong Provincial Key Laboratory of Genome Read and Write (2017B030301011) and Guangdong Provincial Academician Workstation of BGI Synthetic Genomics (2017B090904014). The CKB baseline survey and the first re-survey were supported by a grant from the Kadoorie Charitable Foundation in Hong Kong. The long-term follow-up is supported by grants from the UK Wellcome Trust (212946/Z/18/Z, 202922/Z/16/Z, 104085/Z/14/Z, 088158/Z/09/Z), National Natural Science Foundation of China (81390540, 91846303, 81941018), and Chinese Ministry of Science and Technology (2011BAI09B01). The funders had no role in the study design, data collection, data analysis and interpretation, writing of the report, or the decision to submit the article for publication.

## Data availability

The CKB reference panel and the after-imputed over 100,000 CKB microarray data have been deposited into CNGB Sequence Archive (CNSA) of China National GeneBank DataBase (CNGBdb) with accession number CNP0003405. All genotype data are shared with controlled management.

---

**Figure S1:**
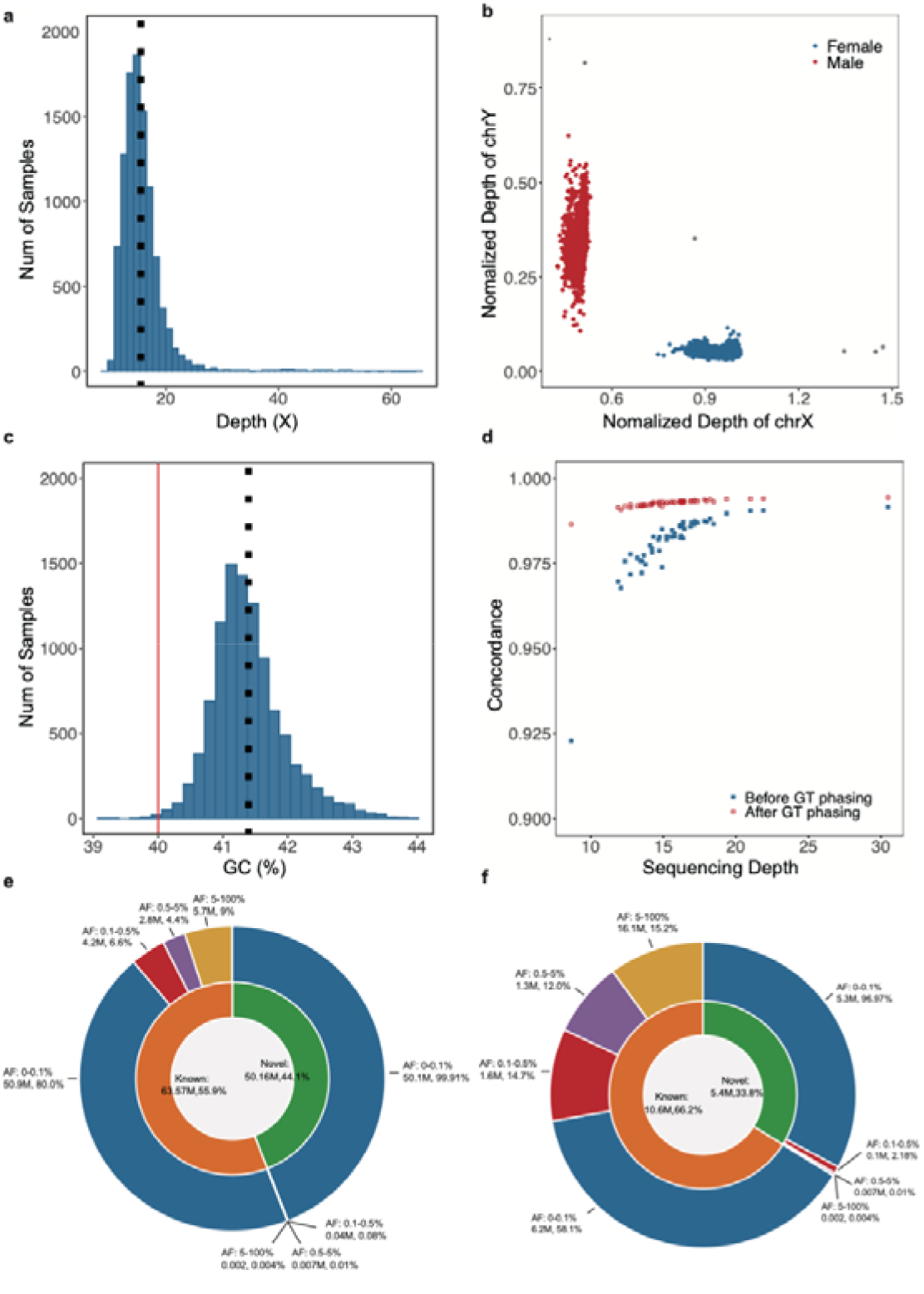
The information about the CKB reference panel. ***Notes*: a**. Distribution of sequencing depth. **b**. The inferred sex of each sample. **c**. Distribution of GC content. **d**. The non-reference concordance rate before and after genotype phasing versus sequencing depth. **e**. The inner circle showed the number and proportion of the novel and known SNPs included in the CKB reference panel. The outer circle showed the number and proportion of SNPs with different AFs. **f**. The inner circle showed the number and proportion of the novel and known INDELs included in the CKB reference panel. The outer circle showed the number and proportion of INDELs with different AFs.

**Figure S2:**
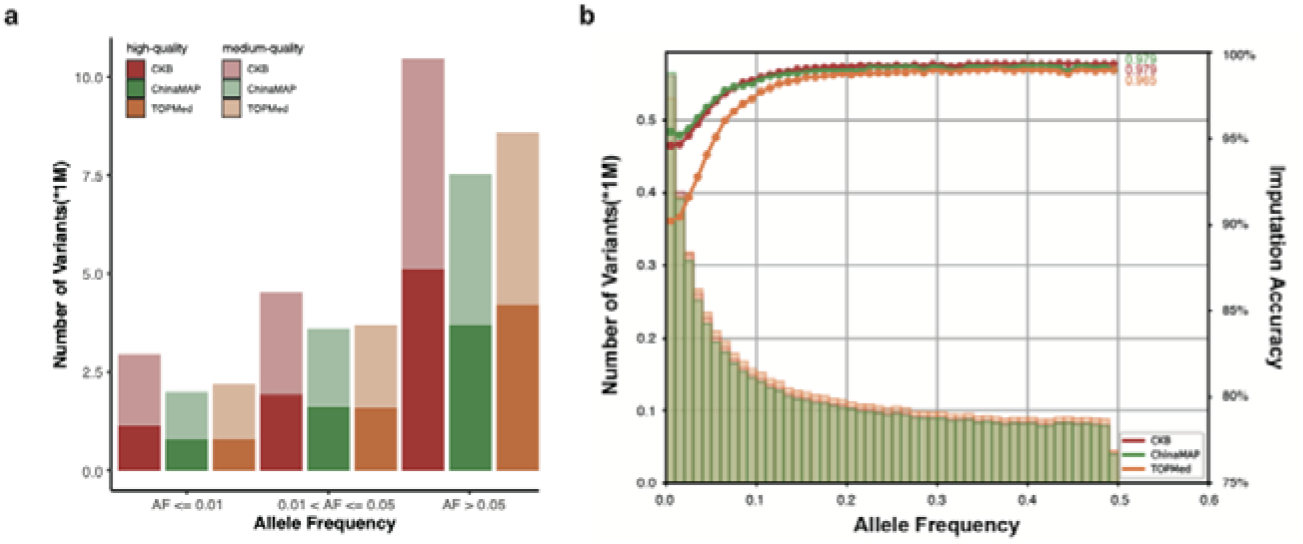
The performance for imputing 1KGP microarray data. ***Notes*: a**. The numbers of high- and medium-quality imputed variants under different AF (allele frequency) by using different reference panels. **b**. The histogram of imputed variants and Pearson correlation coefficients for different panels.

**Figure S3:**
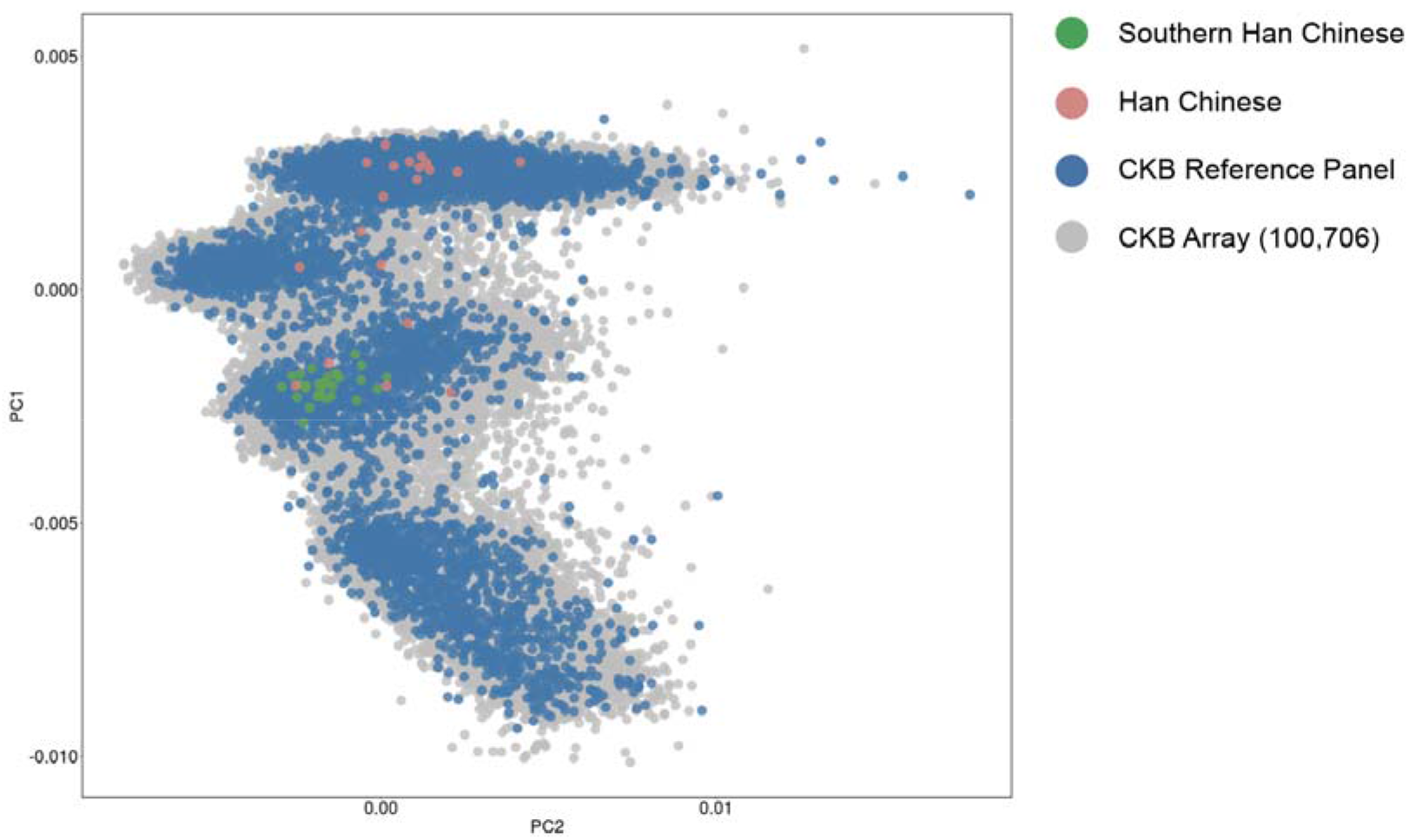
PCA projection of the CKB reference panel and 100,706 CKB microarray data. ***Notes*:** The green, orange, blue, and gray dots represent Southern Han Chinese, Han Chinese, samples in the CKB reference panel, and 100,706 participants in the CKB microarray data.

**Figure S4:**
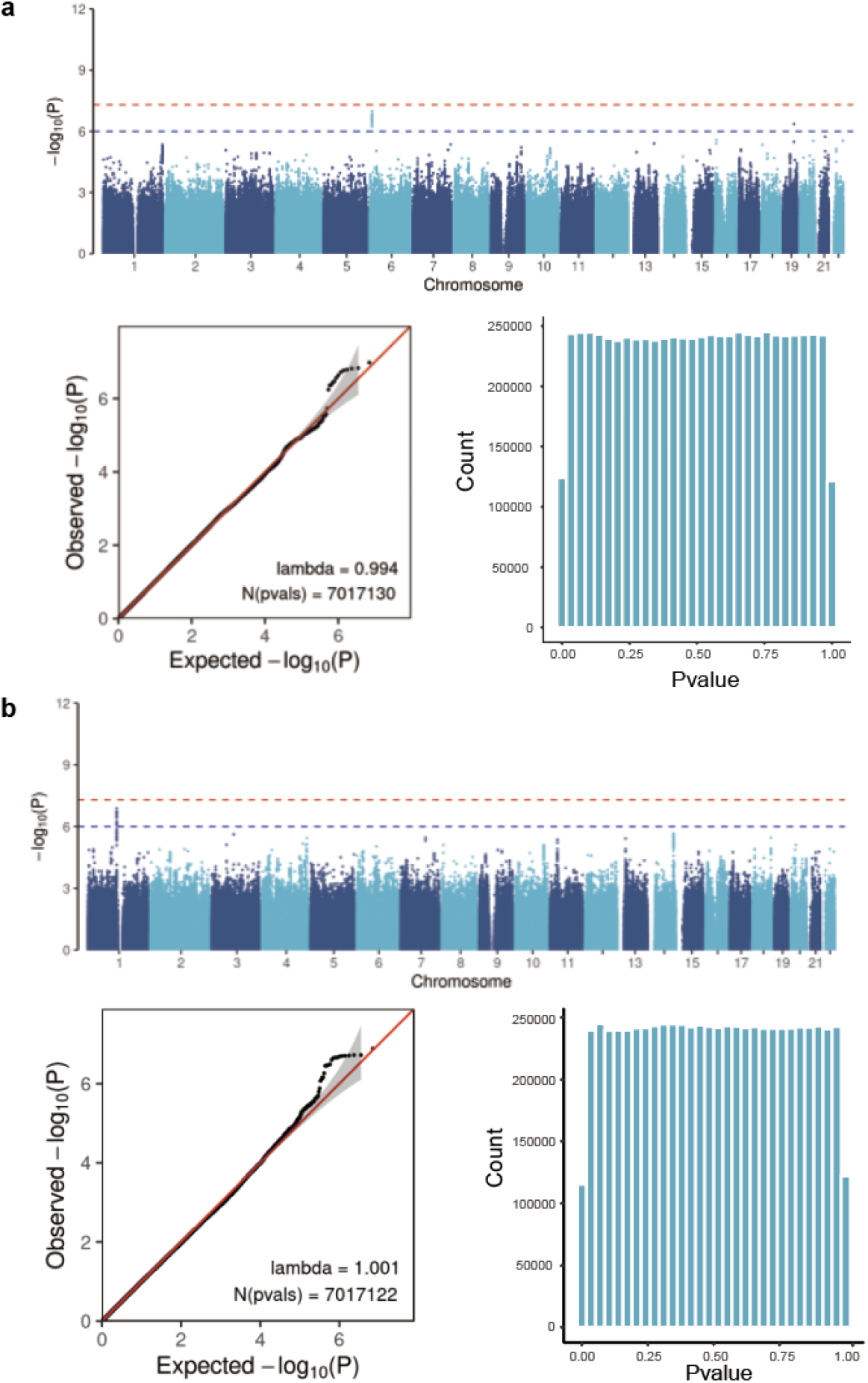
GWAS with simulated phenotypic data based on the CKB reference panel and 100,706 after-imputed CKB microarray data. ***Notes*: a**. The GWAS results with simulated binary phenotypic data under the null hypothesis. **b**. The GWAS results with simulated quantitative phenotypic data under the null hypothesis.

**Figure S5:**
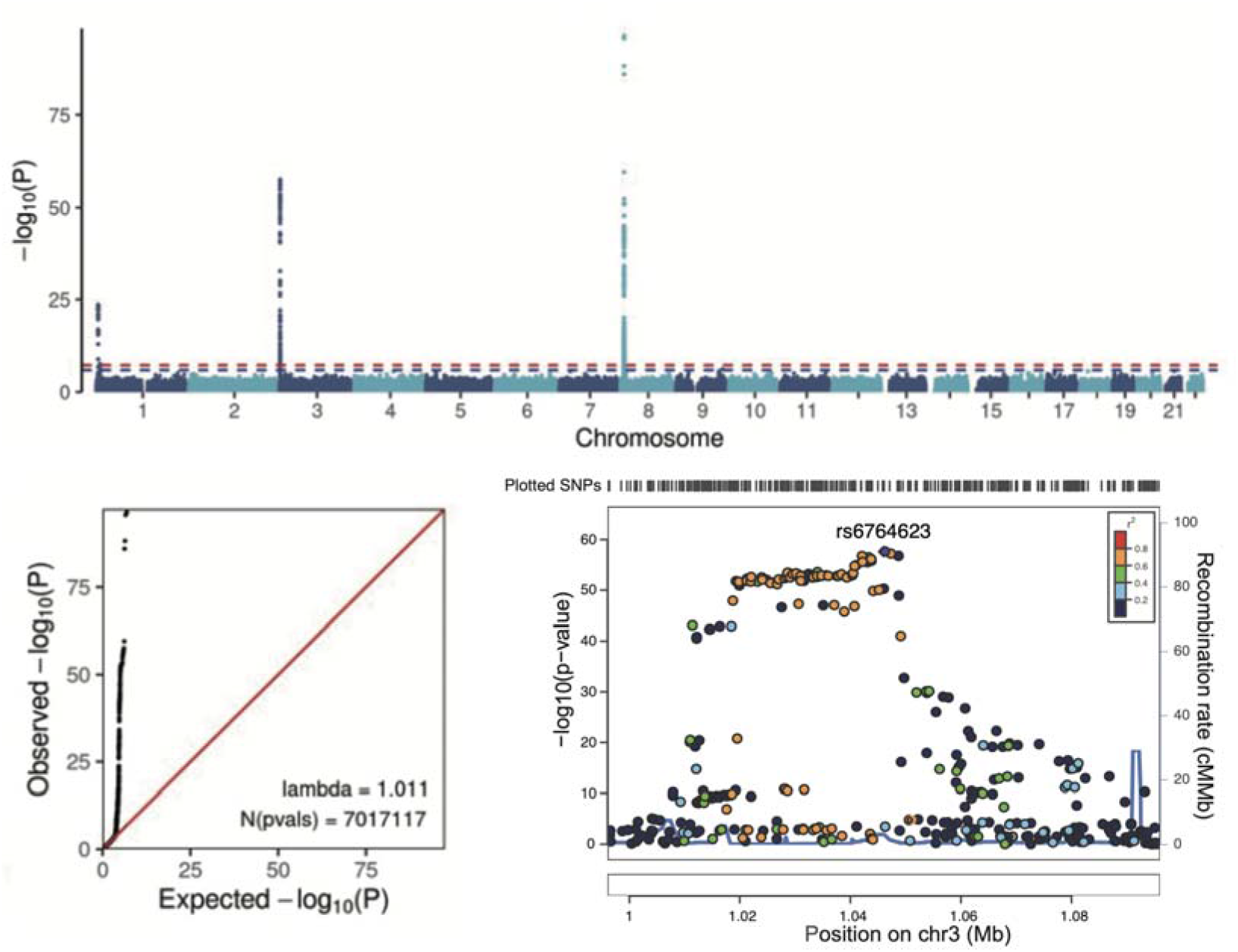
GWAS with simulated phenotypic data based on the CKB reference panel and 100,706 after-imputed CKB microarray data. ***Notes*:** The GWAS results of simulated phenotypic data under the alternative hypothesis.

**Table S1.** The details of the 3×3 confusion matrix.

|  |  | Call state |  |  |
| --- | --- | --- | --- | --- |
|  |  | HOM_REF | HET_REF_VAR | HOM_VAR |
| True state | HOM_REF | TN | FP | FP |
|  | HET_REF_VAR | FN | TP | FP |
|  | HOM_VAR | FN | FP | TP |

**Notes:** HOM\_REF represents homozygous reference allele, HET\_REF\_VAR represents heterozygous (carrying both reference and alternate alleles), and HOM\_VAR represents a homozygous alternate.

**Table S2.** Common Variants were predicted as deleterious by SIFT or PolyPhen.

| CHR | POS | SYMBOL | SIFT | PolyPhen |
| --- | --- | --- | --- | --- |
| 1 | 23484021 | *ASAP3* | Deleterious (0.03) | Possibly_damaging (0.758) |
| 1 | 160195064 | *CASQ1* | Deleterious (0) | Probably_damaging (1) |
| 1 | 204230475 | *PLEKHA6* | Deleterious (0) | Probably_damaging (0.958) |
| 2 | 26187502 | *GAREM2* | Deleterious (0.02) | Benign (0.001) |
| 4 | 139889433 | *MAML3* | Deleterious (0.01) | Probably_damaging (0.994) |
| 4 | 152323001 | *FBXW7* | Deleterious (0.05) | Probably_damaging (0.927) |
| 6 | 158479932 | *TULP4* | Deleterious (0.02) | Probably_damaging (0.93) |
| 7 | 95316772 | *PON1* | Deleterious (0) | Benign (0.031) |
| 9 | 22006132 | *CDKN2B* | Deleterious (0) | Probably_damaging (0.998) |
| 12 | 7652495 | *APOBEC1* | Deleterious (0.03) | Benign (0.299) |
| 14 | 19747851 | *OR4Q3* | Deleterious (0.02) | Benign (0.024) |
| 15 | 28215707 | *HERC2* | Deleterious (0.02) | Benign (0.174) |
| 19 | 57253463 | *ZNF805* | Deleterious (0.02) | Benign (0.281) |
| 22 | 45430916 | *RIBC2* | Deleterious (0.03) | Benign (0.062) |
| 1 | 21814523 | *LDLRAD2* | Tolerated (0.07) | Possibly_damaging (0.694) |
| 1 | 64835430 | *JAK1* | Tolerated (0.17) | Probably_damaging (0.957) |
| 2 | 25744322 | *ASXL2* | Tolerated (0.15) | Probably_damaging (0.984) |
| 2 | 85302574 | *TCF7L1* | Tolerated (0.11) | Possibly_damaging (0.673) |
| 2 | 127719307 | *WDR33* | Tolerated_low_confidence (0.13) | Possibly_damaging (0.514) |
| 5 | 141489460 | *PCDHGC5* | Tolerated (0.24) | Possibly_damaging (0.583) |
| 8 | 29018818 | *HMBOX1* | Tolerated (0.06) | Possibly_damaging (0.861) |
| 10 | 133230794 | *UTF1* | Tolerated (0.11) | Possibly_damaging (0.795) |
| 12 | 120641221 | *CABP1* | Tolerated_low_confidence (0.09) | Possibly_damaging (0.829) |

**Notes:** The 4<sup>th</sup> and 5<sup>th</sup> columns refer to prediction\_type (prediction\_score) by SIFT and PolyPhen, respectively. A SIFT score is a normalized probability of observing the new amino acid at that position, ranging from 0 to 1. A value of 0-0.05 is predicted to affect protein function. For the PolyPhen score, a value close to 1 means likely damaging.

## References

1. Dugger, S.A., A. Platt, and D.B. Goldstein, Drug development in the era of precision medicine. Nature reviews Drug discovery, 2018. 17(3): p. 183–196.

2. Gough, A., et al., Human biomimetic liver microphysiology systems in drug development and precision medicine. Nature Reviews Gastroenterology & Hepatology, 2021. 18(4): p. 252–268.

3. Consortium, I.H., A haplotype map of the human genome. Nature, 2005. 437(7063): p. 1299.

4. Consortium, I.H., A second generation human haplotype map of over 3.1 million SNPs. Nature, 2007. 449(7164): p. 851.

5. Consortium, G.P., A global reference for human genetic variation. Nature, 2015. 526(7571): p. 68.

6. Byrska-Bishop, M., et al., High coverage whole genome sequencing of the expanded 1000 Genomes Project cohort including 602 trios. 2021.

7. McCarthy, S., et al., A reference panel of 64,976 haplotypes for genotype imputation. Nature genetics, 2016. 48(10): p. 1279.

8. Taliun, D., et al., Sequencing of 53,831 diverse genomes from the NHLBI TOPMed Program. Nature, 2021. 590(7845): p. 290–299.

9. Francioli, L.C., et al., Whole-genome sequence variation, population structure and demographic history of the Dutch population. Nature genetics, 2014. 46(8): p. 818–825.

10. Maretty, L., et al., Sequencing and de novo assembly of 150 genomes from Denmark as a population reference. Nature, 2017. 548(7665): p. 87–91.

11. Gudbjartsson, D.F., et al., Large-scale whole-genome sequencing of the Icelandic population. Nature genetics, 2015. 47(5): p. 435–444.

12. Wu, D., et al., Large-scale whole-genome sequencing of three diverse Asian populations in Singapore. Cell, 2019. 179(3): p. 736-749. e15.

13. Cao, Y., et al., The ChinaMAP analytics of deep whole genome sequences in 10,588 individuals. Cell research, 2020. 30(9): p. 717–731.

14. Li, L., et al., The ChinaMAP reference panel for the accurate genotype imputation in Chinese populations. Cell Research, 2021. 31(12): p. 1308–1310.

15. Zhang, P., et al., NyuWa Genome resource: A deep whole-genome sequencing-based variation profile and reference panel for the Chinese population. Cell reports, 2021. 37(7): p. 110017.

16. Chen, Z., et al., Cohort profile: the Kadoorie study of chronic disease in China (KSCDC). International journal of epidemiology, 2005. 34(6): p. 1243–1249.

17. Chen, Z., et al., Contrasting male and female trends in tobacco-attributed mortality in China: evidence from successive nationwide prospective cohort studies. The Lancet, 2015. 386(10002): p. 1447–1456.

18. Bennett, D.A., et al., Association of physical activity with risk of major cardiovascular diseases in Chinese men and women. JAMA cardiology, 2017. 2(12): p. 1349–1358.

19. Du, H., et al., Fresh fruit consumption and major cardiovascular disease in China. N Engl J Med, 2016. 374: p. 1332–1343.

20. Qin, C., et al., Associations of egg consumption with cardiovascular disease in a cohort study of 0.5 million Chinese adults. Heart, 2018. 104(21): p. 1756–1763.

21. Bragg, F., et al., Association between diabetes and cause-specific mortality in rural and urban areas of China. Jama, 2017. 317(3): p. 280–289.

22. Yu, C., et al., Hot tea consumption and its interactions with alcohol and tobacco use on the risk for esophageal cancer: a population-based cohort study. Annals of internal medicine, 2018. 168(7): p. 489–497.

23. Walters, R.G., et al., Genotyping and population structure of the China Kadoorie Biobank. medRxiv, 2022.

24. Wang, J., et al., Genome measures used for quality control are dependent on gene function and ancestry. Bioinformatics, 2015. 31(3): p. 318–323.

25. DePristo, M.A., et al., A framework for variation discovery and genotyping using next-generation DNA sequencing data. Nature genetics, 2011. 43(5): p. 491–498.

26. Browning, B.L., Y. Zhou, and S.R. Browning, A one-penny imputed genome from next-generation reference panels. The American Journal of Human Genetics, 2018. 103(3): p. 338–348.

27. Sherry, S.T., M. Ward, and K. Sirotkin, dbSNP—database for single nucleotide polymorphisms and other classes of minor genetic variation. Genome research, 1999. 9(8): p. 677–679.

28. McCarthy, D.J., et al., Choice of transcripts and software has a large effect on variant annotation. Genome medicine, 2014. 6(3): p. 1–16.

29. Tan, A., G.R. Abecasis, and H.M. Kang, Unified representation of genetic variants. Bioinformatics, 2015. 31(13): p. 2202–2204.

30. Kowalski, M., et al., NHLBI Trans-Omics for Precision Medicine (TOPMed) Consortium; TOPMed Hematology & Hemostasis Working Group: Use of> 100,000 NHLBI Trans-Omics for Precision Medicine (TOPMed) Consortium whole genome sequences improves imputation quality and detection of rare variant associations in admixed African and Hispanic/Latino populations. PLoS Genet, 2019. 15(12): p. e1008500.

31. Zhang, P., et al., NyuWa Genome Resource: Deep Whole Genome Sequencing Based Chinese Population Variation Profile and Reference Panel. bioRxiv, 2021: p. 2020.11. 10.376574.

32. Chen, Y., et al., SOAPnuke: a MapReduce acceleration-supported software for integrated quality control and preprocessing of high-throughput sequencing data. Gigascience, 2018. 7(1): p. gix120.

33. Freed, D., et al., The Sentieon Genomics Tools-A fast and accurate solution to variant calling from next-generation sequence data. BioRxiv, 2017: p. 115717.

34. Jun, G., et al., Detecting and estimating contamination of human DNA samples in sequencing and array-based genotype data. The American Journal of Human Genetics, 2012. 91(5): p. 839–848.

35. Li, H., et al., The sequence alignment/map format and SAMtools. Bioinformatics, 2009. 25(16): p. 2078–2079.

36. Browning, B.L., et al., Fast two-stage phasing of large-scale sequence data. The American Journal of Human Genetics, 2021. 108(10): p. 1880–1890.

37. Manichaikul, A., et al., Robust relationship inference in genome-wide association studies. Bioinformatics, 2010. 26(22): p. 2867–2873.

38. McLaren, W., et al., The ensembl variant effect predictor. Genome biology, 2016. 17(1): p. 1–14.

39. Ng, P.C. and S. Henikoff, SIFT: Predicting amino acid changes that affect protein function. Nucleic acids research, 2003. 31(13): p. 3812–3814.

40. Adzhubei, I.A., et al., A method and server for predicting damaging missense mutations. Nature methods, 2010. 7(4): p. 248–249.

41. Novembre, J. and M. Stephens, Interpreting principal component analyses of spatial population genetic variation. Nature genetics, 2008. 40(5): p. 646–649.

42. Patterson, N., A.L. Price, and D. Reich, Population structure and eigenanalysis. PLoS genetics, 2006. 2(12): p. e190.

43. Purcell, S., et al., PLINK: a tool set for whole-genome association and population-based linkage analyses. The American journal of human genetics, 2007. 81(3): p. 559–575.

44. Chang, C.C., et al., Second-generation PLINK: rising to the challenge of larger and richer datasets. Gigascience, 2015. 4(1): p. s13742-015-0047-8.

45. Fuchsberger, C., G.R. Abecasis, and D.A. Hinds, minimac2: faster genotype imputation. Bioinformatics, 2015. 31(5): p. 782–784.

46. Das, S., et al., Next-generation genotype imputation service and methods. Nature genetics, 2016. 48(10): p. 1284–1287.

